# Causal Modeling of Twitter Activity During COVID-19

**DOI:** 10.1101/2020.05.16.20103903

**Authors:** Oguzhan Gencoglu, Mathias Gruber

## Abstract

Understanding the characteristics of public attention and perception is an essential prerequisite for appropriate crisis management during adverse health events. This is even more crucial during a pandemic such as COVID-19, as primary responsibility of risk management is not centralized to a single institution, but distributed across society. While numerous studies utilize Twitter data in descriptive or predictive context during COVID-19 pandemic, causal modeling of public attention has not been investigated. In this study, we propose a causal inference approach to discover and quantify causal relationships between pandemic characteristics (e.g. number of infections and deaths) and Twitter activity as well as public sentiment. Our results show that the proposed method can successfully capture the epidemiological domain knowledge and identify variables that affect public attention and perception. We believe our work contributes to the field of infodemiology by distinguishing events that correlate with public attention from events that cause public attention.

---

**ON** 11 March 2020, Coronavirus disease 2019 (COVID-19) was declared a pandemic by the World Health Organization [1] and more than 4.4 million people have been infected by it as of 14 May 2020 [2]. During such crises, capturing the dissemination of information, monitoring public opinion, observing compliance to measures, preventing disinformation, and relaying timely information is crucial for risk communication and decision-making about public health [3]. Previous national and global adverse health events show that social media surveillance can be utilized successfully for systematic monitoring of public perception in real-time due to its instantaneous global coverage [4], [5], [6], [7], [8], [9].

Due to its large number of users, Twitter has been the primary social media platform for acquiring, sharing, and spreading information during global adverse events, including the COVID-19 pandemic [10]. Especially during the early stages of the COVID-19 pandemic, millions of posts have been tweeted in a span of couple of weeks by users, i.e., citizens, politicians, corporations, and governmental institutions [11], [12], [13], [14]. Consequently, numerous studies proposed and utilized Twitter as a data source for extracting insights on public health as well as insights on public attention during the COVID-

19 pandemic. Focus of these studies include content analysis [15], topic modeling [16], sentiment analysis [17], nowcasting or forecasting of the disease [18], early detection of the outbreak [19], quantifying and detecting misinformation, disinformation, or conspiracies [20], and measuring public attitude towards relevant health concepts (e.g. social distancing or working from home) [21].

Despite such abundance of studies on manual or automatic analysis of social media data during COVID-19, *causal* modeling of relationships between characteristics of the pandemic and social media activity has not been investigated at all, as of early May 2020. While descriptive statistical analysis (e.g. correlation, cluster, or exploratory analysis) is beneficial for pattern and hypothesis discovery, and standard machine learning methods are effective in predictive modeling of those patterns, causal inference of relevant phenomena will not be possible without causal computational modeling. As adequate assessment of public attention and correct understanding of underlying causes affecting it is imperative for proper decision-making, we hereby propose causal modeling of Twitter activity.

We hypothesize that daily Twitter activity and sentiment during the COVID-19 pandemic has a causal relationship with the characteristics of the pandemic as well as with certain country statistics. We propose a structural causal modeling approach for discovering causal relationships and quantifying likelihood of events under various conditions (i.e. causal queries). To validate our approach, we collect close to 1 million tweets spanning 57 days and identify several attributes of COVID-19 pandemic that might affect Twitter activity. We first employ a structure learning method to automatically construct a graphical causal structure in a data-driven manner. Then, we utilize *Bayesian Networks* (BNs) to learn conditional probability distributions of daily Twitter activity (number of daily tweets) and average public sentiment with respect to several pandemic characteristics such as total number of deaths and number of new infections. Our results show that the proposed structure discovery method can successfully capture the epidemiological domain knowledge. Furthermore, causal inference of daily Twitter activity with cross-validation across 12 countries show that our approach provides accurate predictions of Twitter activity with interpretable and intuitive results. We release the full source code of our study (https://github.com/ogencoglu/causal_twitter_modeling_covid19). We believe our study contributes to the field of infodemiology by proposing causal modeling of public attention during the crisis of COVID-19 pandemic.

## GOING BEYOND CORRELATIONS

Use of observational data from social media was proven to be beneficial in systematic monitoring of public opinion during adverse health events [4], [5], [6], [7], [8], [9]. Such utilization of large, publicly available data becomes even more relevant during a global pandemic such as COVID-19, as neither enough time nor a practical way to run variety of randomized control trials exist. Furthermore, as disease containment measures (e.g. lockdowns, quarantines, and curfews), associated financial issues (e.g. due to inability to work), and changes in social dynamics may impact mental health negatively [22], [23], [24], opinion surveillance methods that do not carry the risk of further stressing of the participants are pertinent. In addition, social media surveillance is less likely to be affected by reporting bias.

Themes of previous studies that focus on exploration of, description of, correlation of, or predictive modeling with Twitter data during COVID-19 pandemic include sentiment analysis [17], [25], [26], [27], [28], public attitude/interest measurement [21], [29], [30], [31], content analysis [32], [33], [15], [34], [35], [36], topic modeling [37], [16], [38], [39], [40], [26], [27], analysis of misinformation, disinformation, or conspiracies [41], [20], [42], [43], [44], [45], [46], outbreak detection or disease nowcasting/forecasting [19], [18], and more [47], [48], [49], [50], [51], [52]. Similarly, data from other social media channels (e.g. Weibo, Reddit, Facebook) or search engine statistics are utilized for parallel analyses related to COVID-19 pandemic as well [53], [54], [55], [56], [57], [58], [59], [60], [61], [62], [63], [64], [65], [66], [67], [68], [69]. While these studies reveal important information and patterns, they do not attempt to uncover or model causal relationships between the attributes of COVID-19 pandemic and social media activity. As *correlation does not imply causation* (e.g. spurious correlations), the ability to identify truly causal relationships between pandemic characteristics and public behaviour (online or not) remains crucial for devising impactful public policies. Without causal understanding, our efforts and decisions on risk communication, public health engagement, health intervention timing, and adjustment of resources for fighting disinformation, fearmongering, and alarmism will stay subpar.

The task of forging causal models comes with numerous challenges in all domains because, typically, domain knowledge and significant amount of time from the experts is required. For substantially complex phenomena such as a pandemic, there are no experts with sufficient understanding to diagnose truly causal attributions. Therefore, learning causal relationships automatically from observational data has been studied in machine learning. One of the primary challenges for this pursuit is that numerous latent variables that we can not observe exist in real world problems. In fact, numerous other latent variables that we are not even aware of may exist as well. As latent variables can induce statistical correlations between observed variables that do not have a causal relationship, *confounding factors* arise. While this phenomenon may not exhibit a considerable problem in standard probabilistic models, causal modeling suffers from it immensely.

Several machine learning methods are proposed for learning causal structures from observational data and some allow combination of statistically significant information (learned from the data) and domain expertise [70], [71]. Bayesian networks are frequently utilized frameworks for learning models once the causal structure is fixed. As probabilistic graphical models, BNs flexibly unify graphical models, structural equations, and counterfactual logic [72], [73], [71], [74]. A causal BN consists of a directed acyclic graph in which nodes correspond to random variables and edges correspond to direct causal influence of one node on another [71]. This compact representation of high-dimensional probability spaces (e.g. joint probability distributions) provides intuitive and explainable models for us. In addition, BNs allow not only straightforward observational computations (e.g. calculation of marginal probabilities) but also interventional ones (e.g. *do-calculus*), enabling simulations of various what-if scenarios.

## METHODS

### Data

We primarily utilized two data sources for our study, i.e., daily number of officially reported COVID-19 infections and deaths from “COVID-19 Data Repository” by the Center for Systems Science and Engineering at Johns Hopkins University [2] and daily count of COVID-19 related tweets from Twitter [75]. A 57 day period between 22 January-18 March 2020 is chosen for this study to represent the early stages of the pandemic when disease characteristics are less known and public panic is elevated. We collected 954,902 tweets from Twitter by searching for *#covid-19* and *#coronavirus* hashtags. Similar to other studies [18], [46], [20], we have extracted the geolocation of the tweets either by using user geo-tagging or geo-coding the information available in users’ profiles, covering 99.2% of the dataset. Timeline of daily log-distribution of collected tweet counts among 177 countries can be examined from **Figure 1**. The trend shows an increasing prevalence of high daily number of tweets as the pandemic spreads across the globe with time.

**Figure 1.**
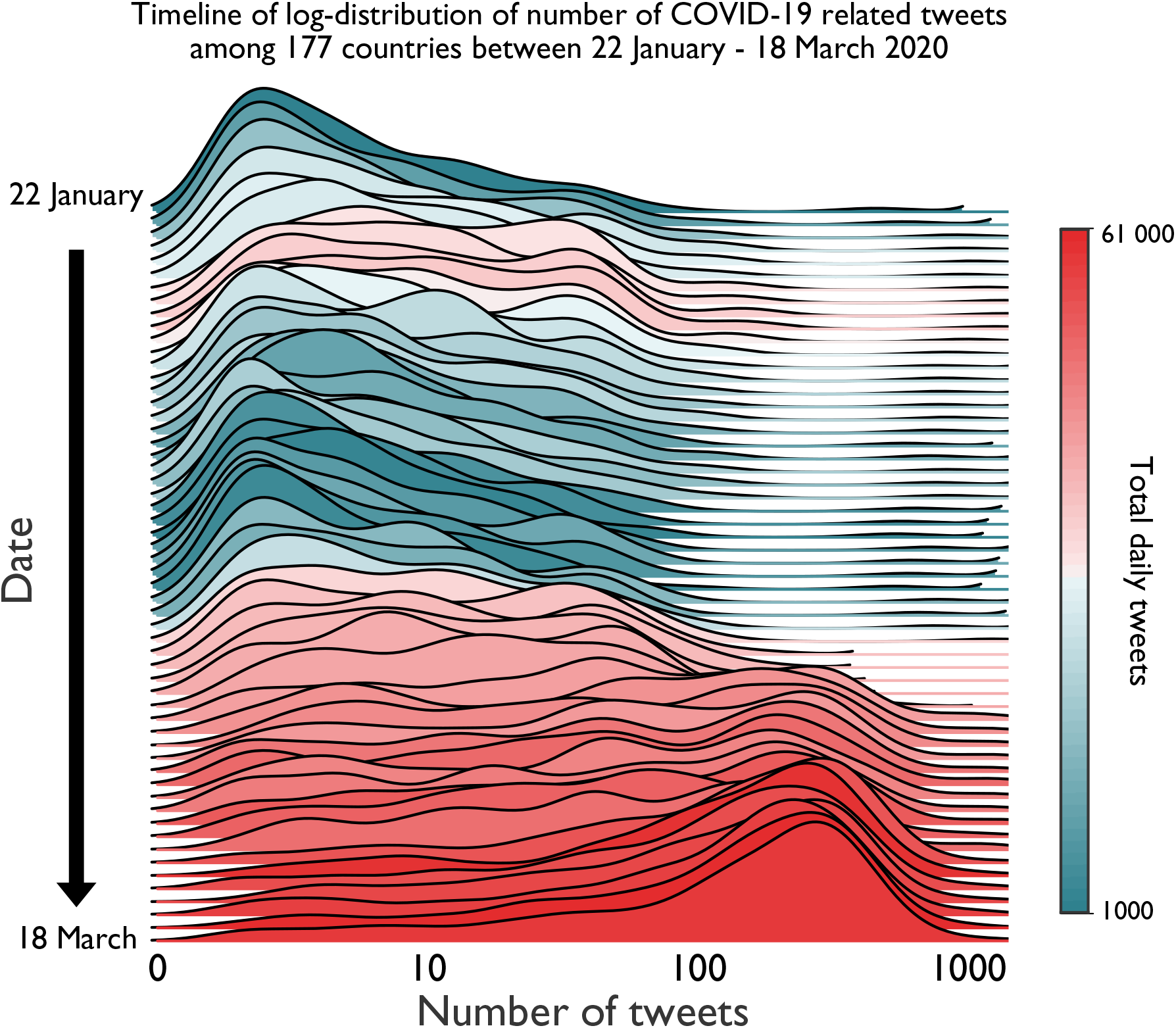
Evolution of COVID-19 related Twitter activity between 22 January – 18 March 2020.

We select the following 12 countries for our causal modeling analysis: Italy, Spain, Germany, France, Switzerland, United Kingdom, Netherlands, Norway, Austria, Belgium, Sweden, and Denmark. These are the countries with substantial number of reported COVID-19 cases (listed in descending order) in Europe as of 18 March 2020, yet still exhibiting a high diversity in terms of the timeline of the pandemic. For instance, while Italy located further in the pandemic timeline due to being hit first in Europe, United Kingdom could be considered in the very initial stages of it for the analysis period of our study. **Figure 2** depicts the cumulative number of tweet counts alongside with that of reported infections and deaths for the selected countries. Evident correlations between these variables can be noticed. A sharp increase in Twitter activity is observed after 28–29 February, which corresponds to the period of each country having at least one confirmed COVID-19 case.

**Figure 2.**
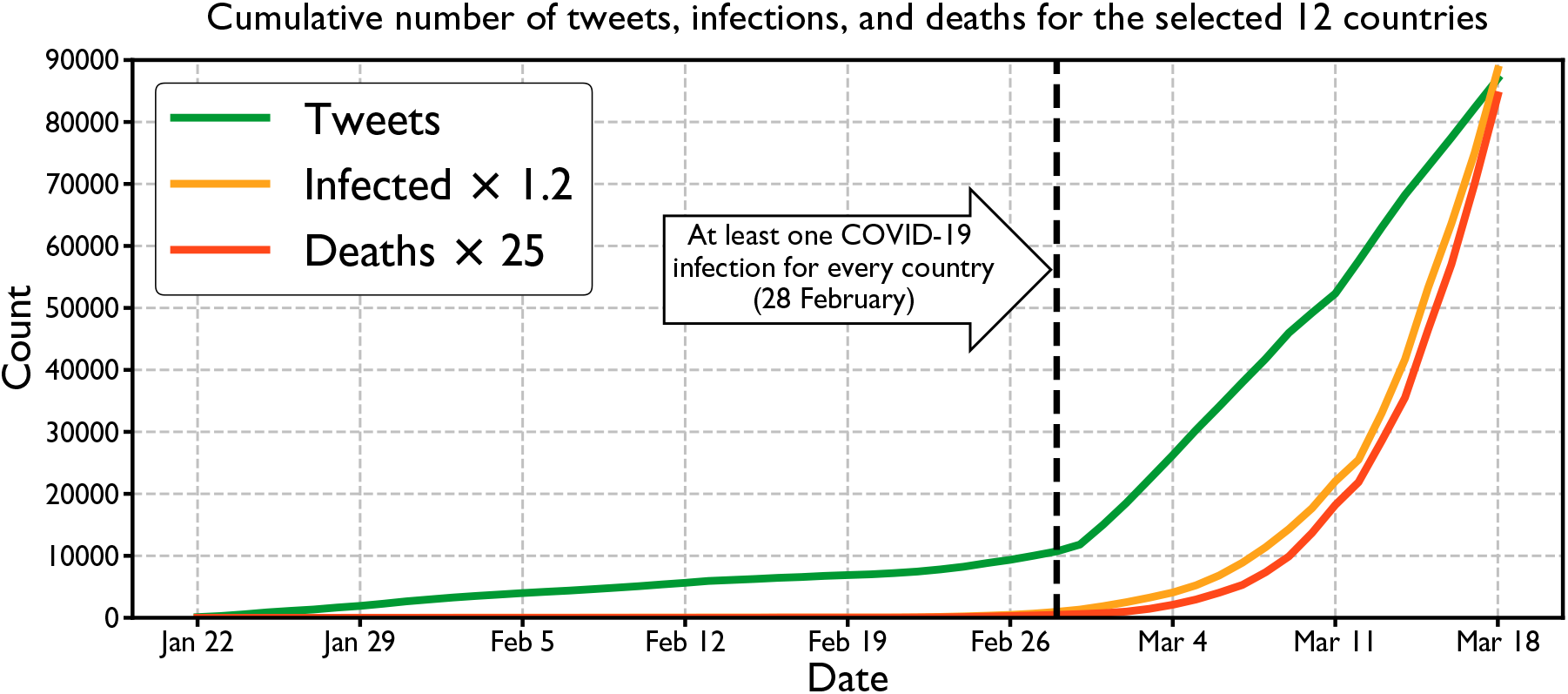
Cumulative counts of Twitter activity and COVID-19 statistics for the selected countries during the study period.

### Feature Selection and Engineering

In order to characterize the pandemic straightforwardly, we calculate the following six features (attributes) from the official COVID-19 incident statistics for each day for 12 selected countries: *(1) total number of infections up to that day* (normalized by the country’s population), (2) *number of new infections* (normalized by the country’s population), (3) *percentage increase in infections* (with respect to previous day), and the same three statistics for *deaths* (4–5–6).

Recent epidemiological studies on COVID-19 reveal the following: people over the age 65 are the primary risk group both for infection and mortality [76], [77], [78], [79] and humanto-human transmission of the virus is largely occurring among family members or among people who co-reside [80], [81], [77]. In order to be able test whether our approach can capture this scientific domain knowledge or not, we collect the following two features for each country: (7) *percentage of population over the age of 65* [82] and (8) *percentage of single-person households* [83]. Finally, as we know that popularity of Twitter in a country and announcement of national lockdown (e.g. closing of schools, banning of gatherings) unequivocally affect the Twitter activity in that country, we add(9) *percentage of population using Twitter* [84] and (10) *is lockdown announced?* (3 day period is encoded as Yes if government restriction is announced [85], No otherwise) features as well. We represent Twitter activity by simply counting the (11) *number of daily tweets* (normalized by the country’s population). We also calculate the (12) *average daily sentiment* (in range [-1, 1]) of English tweets (corresponding to over 80% of all tweets) by utilizing a pre-trained sentiment classifier (DistilBERT [86]). We treat each day as an observation and represent each day with these 12 attributes for structure learning, resulting in a feature matrix of dimensions 684 × 12.

For the purpose of increasing interpretability, we discretize the numerical features by mapping them to 2 categorical levels, namely `High` or `Low`. Features related to the pandemic (infections and deaths) and Twitter activity employ a cutoff value of 75th percentile and remaining numerical features employ a cut-off value of 50th percentile (corresponding to median). Such categorization, for instance, turns the numerical value of “population-normalized increase in deaths of 1.7325 × 10^−7^” into a relatively calculated category of `High` for a given day. Sentiment scores are mapped to `Positive` (≥ 0) or `Negative`(< 0) as well.

### Structure Learning and Causal Inference

We utilize the recently proposed NOTEARS (corresponding to *Non-combinatorial Optimization via Trace Exponential and Augmented lagRangian for Structure learning*) algorithm for structure learning [87]. The algorithm discovers a directed acyclic graph from the observational data by re-formulating the structure learning problem as a purely continuous optimization. This approach differs significantly from existing work in the field which predominantly operates on discrete space of graphs. Consequently, NOTEARS enables utilization of standard numerical solving methods for learning the graph structure in a computationally efficient manner (scales cubically, *𝒪*(*d*^3^), with the number of variables instead of exponentially as in other structure learning methods). As NOTEARS algorithm allows incorporation of expert knowledge, we also put certain constraints on the structure in our experiment. These constraints correspond to prohibited causal attributions based on simple logical assumptions, e.g. Twitter activity on a given day can not have a causal effect on number of deaths from COVID-19 on that day. Once the structure is fixed (both by data and expert knowledge), we treat it as a causal model and learn the parameters of a Bayesian network on it with the training data in order to capture the conditional dependencies between variables. During inference on test data, probabilities of each possible state of a node with respect to the given input data is computed from the conditional probability distributions.

Our approach allows straightforward querying of the model with varying observations. For instance for a given day, the probability of Twitter activity being High, when total number of infections are Low and new deaths are High, i.e.,

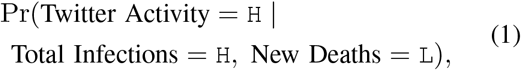

can be computed by propagating the impact of these queries through the nodes of interest. By utilizing this property of our approach, we compute marginal probabilities for gaining further insights on likelihoods of various events.

Essentially, we expect two observations from our experiment. First, we expect the structure learning algorithm to discover the causal relations verified by domain/expert knowledge (e.g. % of single-person households and % of 65+ people affecting infections) and common sense/elementary algebra (e.g. new deaths affecting percentage change in deaths). Second, we expect the calculated likelihoods from the Bayesian network are in parallel with domain knowledge as well, e.g. high % of people over 65 increasing the marginal likelihood of deaths instead of decreasing it or high % of single households (better social isolation) decreasing the marginal likelihood of infections instead of increasing it. Realization of these expectations will show that the proposed method can indeed capture causal relationships and will increase our confidence in discovered relationships between the pandemic attributes and Twitter activity as well as confidence in corresponding likelihoods.

### Evaluation

We validate our approach first by inspecting whether the expected causal relationships (e.g. domain knowledge on COVID-19) are captured or not. Then, we infer the Twitter activity of each day from the learned Bayesian Network. Essentially, this corresponds to a binary classification task, i.e., predicting the Twitter activity as `High` or `Low` from the rest of the variables. We utilize a Leave-One-Country-Out (LOCO) cross-validation scheme in which each fold consists of training set from 11 countries (627 samples) and test set (57 samples) from the remaining country. We do not perform standard kfold cross-validation as we would like to measure the generalization performance across countries and prevent overly optimistic results. Therefore, we ensure that the observations from the same country fall in the same set (either training or test) for every fold. We evaluate the performance of our approach by calculating the average Area Under the Receiver Operating Characteristic curve (AUC) of the cross-validation runs. For quantifying the causal effect of characteristics of pandemic and relevant country statistics on Twitter activity, we report likelihoods from the model by querying various conditions.

## RESULTS

The jointly (with statistical learning from data and user-defined logical constraints) discovered causal model by the structure learning algorithm can be examined from **Figure 3**. Different families of attributes are colored differently for ease of inspection: blue for COVID-19 pandemic related variables, yellow for country-specific statistics, green for government interventions, and red for representing variables related to public attention and perception in Twitter. Daily Twitter activity is affected by four variables, namely Twitter usage statistics of that country, new infections on that day, new deaths on that day, and whether national lockdown is announced or not. Similarly, 4 variables affecting the average daily sentiment in Twitter are new infections on that day, new deaths on that day, total deaths up to that day, and again lockdown announcements. Total number of infections did not show any causal effect on Twitter activity or on average public sentiment.

**Figure 3.**
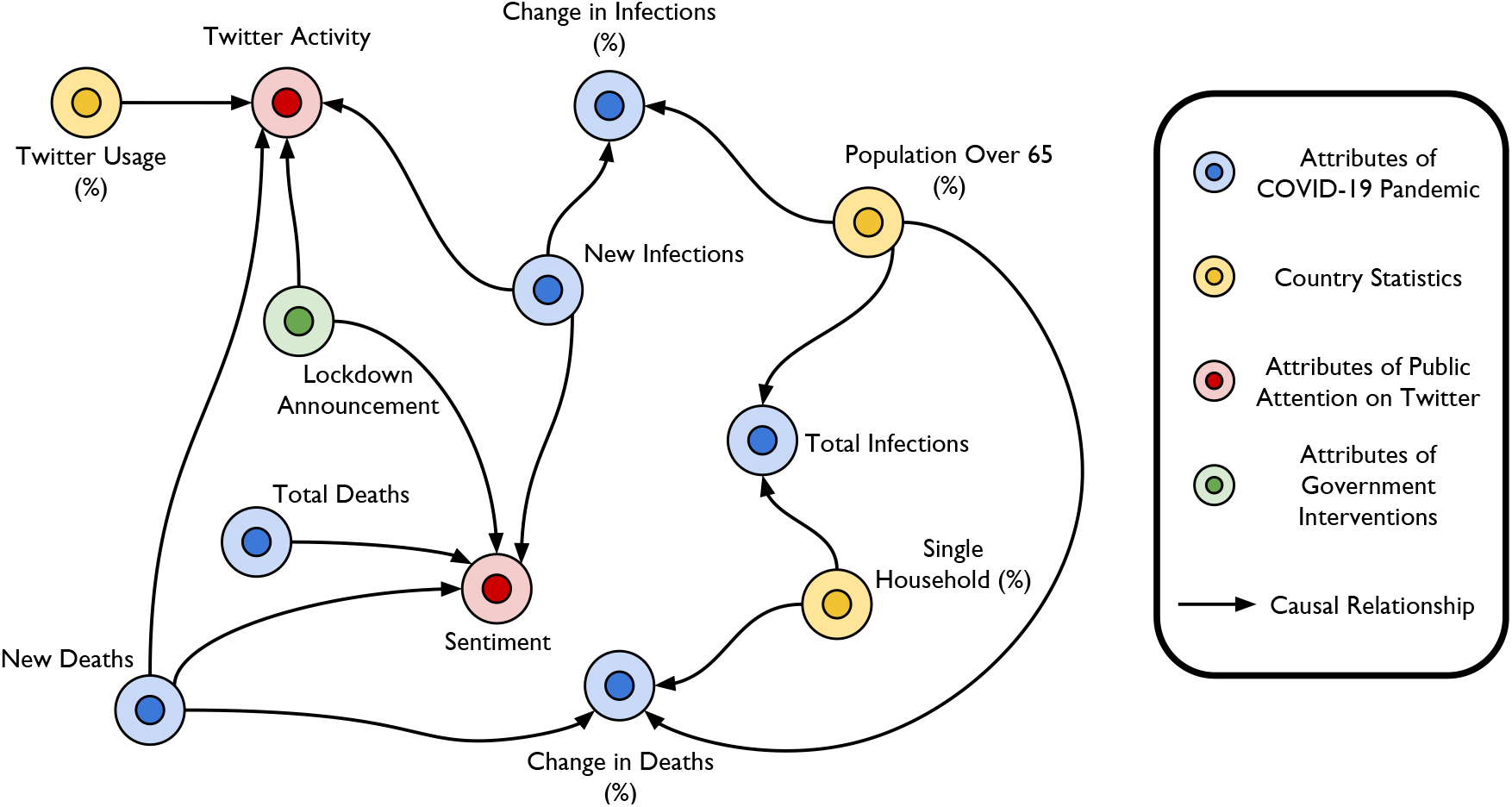
Discovered graph depicting causal relationships between various attributes.

Leave-One-Country-Out cross-validation results in terms of AUCs can be seen in Table 1. Each row in the table corresponds to a crossvalidation fold in which the Twitter activity in that particular country was tried to be predicted. The Bayesian network model achieves an average AUC score of 0.833 across countries when trying to infer the Twitter activity from the rest of the variables for a given day. Daily Twitter patterns of Germany, Italy, and Sweden show very high predictability with AUC scores above 0.97. United Kingdom shows the worst predictability with an AUC of 0.68.

**Table 1.**
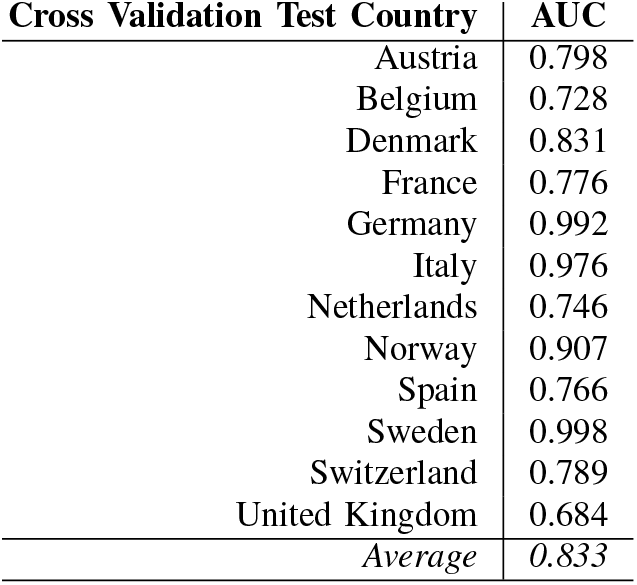
AUC result for each fold of Leave-One-CountryOut cross-validation.

| Cross Validation Test Country | AUC |
| --- | --- |
| Austria | 0.798 |
| Belgium | 0.728 |
| Denmark | 0.831 |
| France | 0.776 |
| Germany | 0.992 |
| Italy | 0.976 |
| Netherlands | 0.746 |
| Norway | 0.907 |
| Spain | 0.766 |
| Sweden | 0.998 |
| Switzerland | 0.789 |
| United Kingdom | 0.684 |
| *Average* | *0.833* |

Calculation of marginal probabilities for several queries are presented in Table 2. Public attention and perception-related target variables and states are set to `High` Twitter Activity and `Negative` Sentiment.

**Table 2.** Examples of queries and computed marginal probabilities for Twitter activity and average sentiment.

| Query | Variable and State | Pr() |
| --- | --- | --- |
| Single-p. hh. (%) = H<br>65+ (%) = L | Total Infections = H | 0.178 |
| Single-p. hh. (%) = L<br>65+ (%) = H | Total Infections = H | 0.241 |
| New Infections = H<br>New Deaths = H | Twitter Activity = H | 0.496 |
| New Infections = L<br>New Deaths = L | Twitter Activity = H | 0.184 |
| New Infections = H<br>New Deaths = H<br>Twitter Usage = H<br>Lockdown Ann. = Yes | Twitter Activity = H | 0.800 |
| New Infections = L<br>New Deaths = L<br>Twitter Usage = L<br>Lockdown Ann. = No | Twitter Activity = H | 0.120 |
| New Deaths = H | Sentiment = Neg | 0.624 |
| New Deaths = L | Sentiment = Neg | 0.277 |
| Total Deaths = H | Sentiment = Neg | 0.344 |
| Total Deaths = L | Sentiment = Neg | 0.290 |
| Lockdown Ann. = Yes | Sentiment = Neg | 0.501 |
| Lockdown Ann. = No | Sentiment = Neg | 0.286 |

## DISCUSSION

By analyzing observational data, we attempt to discover causal associations between national COVID-19 patterns and Twitter activity as well as public sentiment during the early stages of the pandemic. Some of our findings are expected associations such as popularity of Twitter in a country (Twitter usage) affecting Twitter activity. Other expected causal relationships were new deaths affecting change in deaths and new infections affecting change in infections, due to trivial mathematical definitions. These were captured successfully as well. While some of our results imply expected associations, we also observe more interesting implications that are in alignment with recent scientific literature on COVID-19. For instance, percentage of single-person households affects the total number of COVID-19 infections. Similarly, percentage of 65+ population affects the percentage change in deaths (essentially corresponding to rate of deaths). When the queries regarding domain knowledge are examined, we see that low percentage of single-person households (less social isolation) and high percentage of 65+ population increases the probability of total infections being high when compared to the opposite settings. This is in line with recent scientific literature on COVID-19 transmission characteristics [76], [80], [81], [77], [78], [79].

By inferring Twitter activity, we show the generalization ability of causal inference across 12 countries with reasonable accuracy. Factors affecting Twitter activity and sentiment are discussion-worthy as well. By observing correlations, Wong et al. hints that there may be a link between announcement of new infections and Twitter activity [17]. Our results in Figure 3 and Table 2 suggest the same with a causal point of view. Similarly, our finding of negative impact of declaration of government measures on public sentiment is also in parallel with recent research. By analyzing Chinese social media, Li et al. show that official declaration of COVID-19 (epidemic at that time) correlates with increased negative emotions such as anxiety, depression, and indignation [56]. When new infections, new deaths, total deaths are high and an announcement of lockdown is made, Twitter activity on that day becomes more than 6 times more likely than when the situation is opposite (probabilities of 0.8 vs. 0.12). High number of new deaths for a given day causes the sentiment to be much more negative than low number of new deaths (probabilities of 0.624 vs. 0.277). Similarly, an announcement of lockdown is causally associated with an increase in negative sentiment in Twitter (probabilities of 0.501 vs. 0.286).

As it is important to observe the countries that are ahead in terms of pandemic timeline and learn the behaviour of the pandemic, it is equally important to understand also the public perception and behaviour from those countries. Such understanding will aid us in the pursuit of timely decisions and suitable policy-making, and consequently, high public engagement. After all, primary responsibility of risk management during a global pandemic is not centralized to a single institution, but distributed across society. For example, Zhong et al. shows that people’s adherence to COVID-19 control measures is affected by their knowledge and attitudes towards it [88]. In that regard, computational methods such as causal inference and causal reasoning can help us disentangle correlations and causation between the observed variables of the adverse phenomenon.

In real-world scenarios, it is virtually impossible to correctly identify all the causal associations due to presence of numerous confounding factors. As in with all methods in machine learning, a trade-off between false positive associations and false negative ones exists in our approach as well. Furthermore, in the context of this study, ground truth causal associations do not exist even for a few variables, preventing the direct measurement of performance of causal discovery methods. We would like to emphasize that we acknowledge these and other relevant limitations of our study. Our study has further limitations regarding the simplifications on our problem formulation and data. For instance, we do not attempt to model temporal causal relationships in this study, e.g., high deaths numbers having an impact on the public sentiment possibly for several following days. We have not taken into account remarks by famous politicians, public figures, or celebrities which may indeed impact social media discussions. We have not incorporated “retweets” or “likes” into our models either.

Future work includes investigating the effect of dynamics of the pandemic on the spreading mechanisms of information, including relevant health topics in Twitter and other social media. As social media can be exploited for deliberately creating panic and confusion [89], causal inference on patterns of misinformation and disinformation propagation in Twitter will be studied as well. Finally, country-specific models with more granular statistics of the country will be investigated.

## CONCLUSION

Distinguishing epidemiological events that correlate with public attention from epidemiological events that cause public attention is crucial for constructing impactful public health policies. Similarly, monitoring fluctuations of public opinion becomes actionable only if causal relationships are identified. We hope our study serves as a first example of causal inference on social media data for increasing our understanding of factors affecting public attention and perception during COVID-19 pandemic.

## Data Availability

The data of this study is available at https://github.com/ogencoglu/causal_twitter_modeling_covid19

## reference

1. D. Cucinotta and M. Vanelli, “Who declares covid-19 a pandemic.” Acta Bio-medica: Atenei Parmensis, vol. 91, no. 1, pp. 157–160, 2020.

2. E. Dong, H. Du, and L. Gardner, “An interactive web-based dashboard to track covid-19 in real time,” The Lancet Infectious Diseases, 2020.

3. J.J. Van Bavel, K. Baicker, P.S. Boggio, V. Capraro, A. Cichocka, M. Cikara, M.J. Crockett, A.J. Crum, K.M. Douglas, J.N. Druckman et al., “Using social and behavioural science to support covid-19 pandemic response,” Nature Human Behaviour, pp. 1–12, 2020.

4. A. Signorini, A.M. Segre, and P.M. Polgreen, “The use of twitter to track levels of disease activity and public concern in the us during the influenza a h1n1 pandemic,” PloS One, vol. 6, no. 5, 2011.

5. X. Ji, S.A. Chun, and J. Geller, “Monitoring public health concerns using twitter sentiment classifications,” in IEEE International Conference on Healthcare Informatics. IEEE, 2013, pp. 335–344.

6. X. Ji, S.A. Chun, Z. Wei, and J. Geller, “Twitter sentiment classification for measuring public health concerns,” Social Network Analysis and Mining, vol. 5, no. 1, p. 13, 2015.

7. C. Weeg, H.A. Schwartz, S. Hill, R.M. Merchant, C. Arango, and L. Ungar, “Using twitter to measure public discussion of diseases: a case study,” JMIR Public Health and Surveillance, vol. 1, no. 1, p. e6, 2015.

8. L. Mollema, I.A. Harmsen, E. Broekhuizen, R. Clijnk, H. De Melker, T. Paulussen, G. Kok, R. Ruiter, and E. Das, “Disease detection or public opinion reflection? content analysis of tweets, other social media, and online newspapers during the measles outbreak in the netherlands in 2013,” Journal of Medical Internet Research (JMIR), vol. 17, no. 5, p. e128, 2015.

9. S.E. Jordan, S.E. Hovet, I.C.-H. Fung, H. Liang, K.-W. Fu, and Z.T.H. Tse, “Using twitter for public health surveillance from monitoring and prediction to public response,” Data, vol. 4, no. 1, p. 6, 2019.

10. H. Rosenberg, S. Syed, and S. Rezaie, “The twitter pandemic: the critical role of twitter in the dissemination of medical information and misinformation during the covid-19 pandemic,” Canadian Journal of Emergency Medicine, pp. 1–7x, 2020.

11. E. Chen, K. Lerman, and E. Ferrara, “Covid-19: The first public coronavirus twitter dataset,” arXiv preprint arXiv:2003.07372, 2020.

12. Z. Gao, S. Yada, S. Wakamiya, and E. Aramaki, “Naist covid: Multilingual covid-19 twitter and weibo dataset,” arXiv preprint arXiv:2004.08145, 2020.

13. R. Lamsal, “Corona virus (covid-19) tweets dataset,” 2020. [Online]. Available: http://dx.doi.org/10.21227/781w-ef42

14. N. Aguilar-Gallegos, L.E. Romero-García, E.G. Martínez-Gonzaález, E.I. Garcaí-Sánchez, and J. Aguilar-Ávila, “Dataset on dynamics of coronavirus on twitter,” Data in Brief, p. 105684, 2020.

15. M. Thelwall and S. Thelwall, “Retweeting for covid-19: Consensus building, information sharing, dissent, and lockdown life,” arXiv preprint arXiv:2004.02793, 2020.

16. H. Sha, M.A. Hasan, G. Mohler, and P.J. Brantingham, “Dynamic topic modeling of the covid-19 twitter narrative among us governors and cabinet executives,” arXiv preprint arXiv:2004.11692, 2020.

17. C.M.L. Wong and O. Jensen, “The paradox of trust: perceived risk and public compliance during the covid-19 pandemic in singapore,” Journal of Risk Research, pp. 1–10, 2020.

18. J. Turiel and T. Aste, “Wisdom of the crowds in forecasting covid-19 spreading severity,” arXiv preprint arXiv:2004.04125, 2020.

19. E. Gharavi, N. Nazemi, and F. Dadgostari, “Early outbreak detection for proactive crisis management using twitter data: Covid-19 a case study in the us,” arXiv preprint arXiv:2005.00475, 2020.

20. M. Chary, D. Overbeek, A. Papadimoulis, A. Sheroff, and M. Burns, “Geospatial correlation between covid-19 health misinformation on social media and poisoning with household cleaners,” medRxiv, 2020.

21. A. Kayes, M.S. Islam, P.A. Watters, A. Ng, and H. Kayesh, “Automated measurement of attitudes towards social distancing using social media: A covid-19 case study,” Preprints, 2020.

22. C. Wang, R. Pan, X. Wan, Y. Tan, L. Xu, C.S. Ho, and R.C. Ho, “Immediate psychological responses and associated factors during the initial stage of the 2019 coronavirus disease (covid-19) epidemic among the general population in china,” International Journal of Environmental Research and Public Health, vol. 17, no. 5, p. 1729, 2020.

23. W. Cullen, G. Gulati, and B. Kelly, “Mental health in the covid-19 pandemic,” QJM: An International Journal of Medicine, vol. 113, no. 5, pp. 311–312, 2020.

24. S.K. Brooks, R.K. Webster, L.E. Smith, L. Woodland, S. Wessely, N. Greenberg, and G.J. Rubin, “The psychological impact of quarantine and how to reduce it: rapid review of the evidence,” The Lancet, 2020.

25. A.D. Dubey and S. Tripathi, “Analysing the sentiments towards work-from-home experience during covid-19 pandemic,” Journal of Innovation Management, vol. 8, no. 1, 2020.

26. V. Duong, P. Pham, T. Yang, Y. Wang, and J. Luo, “The ivory tower lost: How college students respond differently than the general public to the covid-19 pandemic,” arXiv preprint arXiv:2004.09968, 2020.

27. R.J. Medford, S.N. Saleh, A. Sumarsono, T.M. Perl, and C.U. Lehmann, “An” infodemic”: Leveraging high-volume twitter data to understand public sentiment for the covid-19 outbreak,” medRxiv, 2020.

28. J. Samuel, G.M.N. Ali, M. M. Rahman, E. Esawi, and Y. Samuel, “Covid-19 public sentiment insights and machine learning for tweets classification,” Preprints, 2020.

29. Z. Batooli and M. Sayyah, “Measuring social media attention of scientific research on novel coronavirus disease 2019 (covid-19): An investigation on article-level metrics data of dimensions,” Preprint from Research Square, 2020.

30. J. Kwon, C. Grady, J.T. Feliciano, and S.J. Fodeh, “Defining facets of social distancing during the covid-19 pandemic: Twitter analysis,” medRxiv, 2020.

31. M. Cinelli, W. Quattrociocchi, A. Galeazzi, C.M. Valensise, E. Brugnoli, A.L. Schmidt, P. Zola, F. Zollo, and A. Scala, “The covid-19 social media infodemic,” arXiv preprint arXiv:2003.05004, 2020.

32. H.W. Park, S. Park, and M. Chong, “Conversations and medical news frames on twitter: Infodemiological study on covid-19 in south korea,” Journal of Medical Internet Research (JMIR), vol. 22, no. 5, p. e18897, 2020.

33. M. Thelwall and S. Thelwall, “Covid-19 tweeting in english: Gender differences,” arXiv preprint arXiv:2003.11090, 2020.

34. T. Alshaabi, J. Minot, M. Arnold, J.L. Adams, D.R. Dewhurst, A.J. Reagan, R. Muhamad, C.M. Danforth, and P.S. Dodds, “How the world’s collective attention is being paid to a pandemic: Covid-19 related 1-gram time series for 24 languages on twitter,” arXiv preprint arXiv:2003.12614, 2020.

35. C.E. Lopez, M. Vasu, and C. Gallemore, “Understanding the perception of covid-19 policies by mining a multilanguage twitter dataset,” arXiv preprint arXiv:2003.10359, 2020.

36. D.R. Dewhurst, T. Alshaabi, M.V. Arnold, J.R. Minot, C.M. Danforth, and P.S. Dodds, “Divergent modes of online collective attention to the covid-19 pandemic are associated with future caseload variance,” arXiv preprint arXiv:2004.03516, 2020.

37. A. Abd-Alrazaq, D. Alhuwail, M. Househ, M. Hamdi, and Z. Shah, “Top concerns of tweeters during the covid-19 pandemic: infoveillance study,” Journal of Medical Internet Research (JMIR), vol. 22, no. 4, p. e19016, 2020.

38. P. Wicke and M. M. Bolognesi, “Framing covid-19: How we conceptualize and discuss the pandemic on twitter,” arXiv preprint arXiv:2004.06986, 2020.

39. A. Jarynowski, M. Woójta-Kempa, and V. Belik, “Trends in perception of covid-19 in polish internet,” medRxiv, 2020.

40. C. Ordun, S. Purushotham, and E. Raff, “Exploratory analysis of covid-19 tweets using topic modeling, umap, and digraphs,” arXiv preprint arXiv:2005.03082, 2020.

41. K.-C. Yang, C. Torres-Lugo, and F. Menczer, “Prevalence of low-credibility information on twitter during the covid-19 outbreak,” arXiv preprint arXiv:2004.14484, 2020.

42. W. Ahmed, J. Vidal-Alaball, J. Downing, and F.L. Seguí, “Covid-19 and the 5g conspiracy theory: Social network analysis of twitter data,” Journal of Medical Internet Research (JMIR), vol. 22, no. 5, p.e19458, 2020.

43. E. Ferrara, “#covid-19 on twitter: Bots, conspiracies, and social media activism,” arXiv preprint arXiv:2004.09531, 2020.

44. A. Bridgman, E. Merkley, P.J. Loewen, T. Owen, D. Ruths, L. Teichmann, and O. Zhilin, “The causes and consequences of covid-19 misperceptions: Understanding the role of news and social media,” OSF Preprints, 2020.

45. W. Ahmed, J. Vidal-Alaball, J. Downing, and F.L. Seguí, “Dangerous messages or satire? analysing the conspiracy theory linking 5g to covid-19 through social network analysis,” Journal of Medical Internet Research (JMIR), 2020.

46. R. Gallotti, F. Valle, N. Castaldo, P. Sacco, and M. De Domenico, “Assessing the risks of” infodemics” in response to covid-19 epidemics,” arXiv preprint arXiv:2004.03997, 2020.

47. S. Golder, A. Klein, A. Magge, K. O’Connor, H. Cai, and D. Weissenbacher, “Extending a chronological and geographical analysis of personal reports of covid-19 on twitter to england, uk,” medRxiv, 2020.

48. A. Sarker, S. Lakamana, W. Hogg-Bremer, A. Xie, M.A. Al-Garadi, and Y.-C. Yang, “Self-reported covid-19 symptoms on twitter: An analysis and a research resource,” medRxiv, 2020.

49. I. Li, Y. Li, T. Li, S. Alvarez-Napagao, and D. Garcia, “What are we depressed about when we talk about covid-19: Mental health analysis on tweets using natural language processing,” arXiv preprint arXiv:2004.10899, 2020.

50. P. Xu, M. Dredze, and D.A. Broniatowski, “The twitter social mobility index: Measuring social distancing practices from geolocated tweets,” arXiv preprint arXiv:2004.02397, 2020.

51. H. Lyu, L. Chen, Y. Wang, and J. Luo, “Sense and sensibility: Characterizing social media users regarding the use of controversial terms for covid-19,” arXiv preprint arXiv:2004.06307, 2020.

52. L. Schild, C. Ling, J. Blackburn, G. Stringhini, Y. Zhang, and S. Zannettou, “” go eat a bat, chang!”: An early look on the emergence of sinophobic behavior on web communities in the face of covid-19,” arXiv preprint arXiv:2004.04046, 2020.

53. A. Rovetta and A. S. Bhagavathula, “Covid-19-related web search behaviors and infodemic attitudes in italy: Infodemiological study,” JMIR Public Health and Surveillance, vol. 6, no. 2, p. e19374, 2020.

54. S. Shahsavari, P. Holur, T.R. Tangherlini, and V. Roychowdhury, “Conspiracy in the time of corona: Automatic detection of covid-19 conspiracy theories in social media and the news,” arXiv preprint arXiv:2004.13783, 2020.

55. J. Li, Q. Xu, R. Cuomo, V. Purushothaman, and T. Mackey, “Data mining and content analysis of the chinese social media platform weibo during the early covid-19 outbreak: retrospective observational infoveillance study,” JMIR Public Health and Surveillance, vol. 6, no. 2, p.e18700, 2020.

56. S. Li, Y. Wang, J. Xue, N. Zhao, and T. Zhu, “The impact of covid-19 epidemic declaration on psychological consequences: A study on active weibo users,” International Journal of Environmental Research and Public Health, vol. 17, no. 6, p. 2032, 2020.

57. N. Velaásquez, R. Leahy, N.J. Restrepo, Y. Lupu, R. Sear, N. Gabriel, O. Jha, and N. Johnson, “Hate multiverse spreads malicious covid-19 content online beyond individual platform control,” arXiv preprint arXiv:2004.00673, 2020.

58. Y. Zhao and H. Xu, “Chinese public attention to covid-19 epidemic: Based on social media,” medRxiv, 2020.

59. L. Li, Q. Zhang, X. Wang, J. Zhang, T. Wang, T.-L. Gao, W. Duan, K. K.-f. Tsoi, and F.-Y. Wang, “Characterizing the propagation of situational information in social media during covid-19 epidemic: A case study on weibo,” IEEE Transactions on Computational Social Systems, vol. 7, no. 2, pp. 556–562, 2020.

60. V. Lampos, S. Moura, E. Yom-Tov, I.J. Cox, R. McKendry, and M. Edelstein, “Tracking covid-19 using online search,” arXiv preprint arXiv:2003.08086, 2020.

61. S. Boberg, T. Quandt, T. Schatto-Eckrodt, and L. Frischlich, “Pandemic populism: Facebook pages of alternative news media and the corona crisis– a computational content analysis,” arXiv preprint arXiv:2004.02566, 2020.

62. H. Jelodar, Y. Wang, R. Orji, and H. Huang, “Deep sentiment classification and topic discovery on novel coronavirus or covid-19 online discussions: Nlp using lstm recurrent neural network approach,” arXiv preprint arXiv:2004.11695, 2020.

63. D. Liu, L. Clemente, C. Poirier, X. Ding, M. Chinazzi, J.T. Davis, A. Vespignani, and M. Santillana, “A machine learning methodology for real-time forecasting of the 2019–2020 covid-19 outbreak using internet searches, news alerts, and estimates from mechanistic models,” arXiv preprint arXiv:2004.04019, 2020.

64. Z. Hou, F. Du, H. Jiang, X. Zhou, and L. Lin, “Assessment of public attention, risk perception, emotional and behavioural responses to the covid-19 outbreak: social media surveillance in china,” medRxiv preprint, 2020.

65. D.C. Stokes, A. Andy, S.C. Guntuku, L.H. Ungar, and R.M. Merchant, “Public priorities and concerns regarding covid-19 in an online discussion forum: Longitudinal topic modeling,” Journal of General Internal Medicine, 2020.

66. C. Shen, A. Chen, C. Luo, W. Liao, J. Zhang, and B. Feng, “Reports of own and others’ symptoms and diagnosis on social media predict covid-19 case counts in mainland china,” arXiv preprint arXiv:2004.06169, 2020.

67. Q. Chen, C. Min, W. Zhang, G. Wang, X. Ma, and R. Evans, “Unpacking the black box: How to promote citizen engagement through government social media during the covid-19 crisis,” Computers in Human Behavior, p. 106380, 2020.

68. B. Lucas, B. Elliot, and T. Landman, “Online information search during covid-19,” arXiv preprint arXiv:2004.07183, 2020.

69. E.A. Pekoz, A. Smith, A. Tucker, and Z. Zheng, “Covid19 symptom web search surges precede local hospitalization surges,” Available at SSRN 3585532, 2020.

70. B. Ellis and W. H. Wong, “Learning causal bayesian network structures from experimental data,” Journal of the American Statistical Association, vol. 103, no. 482, pp. 778–789, 2008.

71. D. Koller and N. Friedman, Probabilistic graphical models: principles and techniques. MIT press, 2009.

72. D.B. Rubin, “Causal inference using potential outcomes: Design, modeling, decisions,” Journal of the American Statistical Association, vol. 100, no. 469, pp. 322–331, 2005.

73. J. Pearl, “An introduction to causal inference,” The International Journal of Biostatistics, vol. 6, no. 2, 2010.

74. J. Pearl, Causality. Cambridge University Press, 2009.

75. “Twitter,” https://twitter.com/, accessed: 2020-05-12.

76. J.B. Dowd, V. Rotondi, L. Adriano, D.M. Brazel, P. Block, X. Ding, Y. Liu, and M.C. Mills, “Demographic science aids in understanding the spread and fatality rates of covid-19,” medRxiv, 2020.

77. Y.-R. Guo, Q.-D. Cao, Z.-S. Hong, Y.-Y. Tan, S.-D. Chen, H.-J. Jin, K.-S. Tan, D.-Y. Wang, and Y. Yan, “The origin, transmission and clinical therapies on coronavirus disease 2019 (covid-19) outbreak–an update on the status,” Military Medical Research, vol. 7, no. 1, pp. 1– 10, 2020.

78. X. Yang, Y. Yu, J. Xu, H. Shu, H. Liu, Y. Wu, L. Zhang, Z. Yu, M. Fang, T. Yu et al., “Clinical course and outcomes of critically ill patients with sars-cov-2 pneumonia in wuhan, china: a single-centered, retrospective, observational study,” The Lancet Respiratory Medicine, 2020.

79. W. Wang, J. Tang, and F. Wei, “Updated understanding of the outbreak of 2019 novel coronavirus (2019-ncov) in wuhan, china,” Journal of Medical Virology, vol. 92, no. 4, pp. 441–447, 2020.

80. WHO, “Report of the who-china joint mission on coronavirus disease 2019 (covid-19),” Geneva: World Health Organization, 2020.

81. C. Li, F. Ji, L. Wang, J. Hao, M. Dai, Y. Liu, X. Pan, J. Fu, L. Li, G. Yang et al., “Asymptomatic and human-to-human transmission of sars-cov-2 in a 2-family cluster, xuzhou, china.” Emerging Infectious Diseases, vol. 26, no. 7, 2020.

82. “World bank open data – population ages 65 and above,” https://data.worldbank.org/, accessed: 2020-05-12.

83. “Distribution of households by household type from 2003 onwards – eu-silc survey,” https://appsso.eurostat.ec.europa.eu/nui/show.do?dataset=ilclvph02&lang=en, accessed: 2020-05-12.

84. “Social media stats – february 2020,” https://gs.statcounter.com/ accessed: 2020-05-12.

85. “National responses to the covid-19 pandemic – lockdown data,” https://en.wikipedia.org/wiki/National_responses_to_the_COVID-19_pandemic, accessed: 2020-05-12.

86. V. Sanh, L. Debut, J. Chaumond, and T. Wolf, “Distilbert, a distilled version of bert: smaller, faster, cheaper and lighter,” arXiv preprint arXiv:1910.01108, 2019.

87. X. Zheng, B. Aragam, P.K. Ravikumar, and E.P. Xing, “Dags with no tears: Continuous optimization for structure learning,” in Advances in Neural Information Processing Systems, 2018, pp. 9472–9483.

88. B.-L. Zhong, W. Luo, H.-M. Li, Q.-Q., Zhang, X.-G. Liu, W.-T. Li, and Y. Li, “Knowledge, attitudes, and practices towards covid-19 among chinese residents during the rapid rise period of the covid-19 outbreak: a quick online cross-sectional survey,” International Journal of Biological Sciences, vol. 16, no. 10, p. 1745, 2020.

89. R.M. Merchant and N. Lurie, “Social media and emergency preparedness in response to novel coronavirus,” Journal of the American Medical Association (JAMA), 2020.

